# Anti-SARS-CoV-2 antibodies seroprevalence among patients submitted to treatment for tuberculosis in Rio de Janeiro, Brazil: a cross-sectional study

**DOI:** 10.1101/2021.11.17.21266274

**Authors:** Karen Machado Gomes, Suzanne Pereira Leite, Maria Helena Vieira Moutinho, Thatiana Alfena de Souza, Rita de Cássia Batista, Luiz Claudio Motta de Oliveira, Paulo Redner, Jesus Pais Ramos, Fatima Maria Gomes da Rocha, Gisele Pinto de Oliveira, Antônio Teva, Fernando do Couto Motta, Marilda Agudo Mendonça Teixeira de Siqueira, Rafael Silva Duarte, Francisco Inácio Pinkusfeld Monteiro Bastos, Paulo Victor de Sousa Viana

**Affiliations:** Centro de Referência Professor Helio Fraga, Escola Nacional de Saúde Pública Sergio Arouca, Fundação Oswaldo Cruz – Campus Jacarepaguá, Estrada de Curicica, 2000 - Curicica, Rio de Janeiro - RJ, 22780-195, Brazil; Vice-Diretoria de Ambulatório e Laboratório, Escola Nacional de Saúde Pública Sergio Arouca, Fundação Oswaldo Cruz, Av. Brasil, 4365 - Manguinhos, Rio de Janeiro - RJ, 21040-900, Brazil; Departamento de Ciências Biológicas, Escola Nacional de Saúde Pública Sergio Arouca, Fundação Oswaldo Cruz, Av. Brasil, 4365 - Manguinhos, Rio de Janeiro - RJ, 21040-900, Brazil; Laboratório de Vírus Respiratórios e Sarampo, Instituto Oswaldo Cruz, Fundação Oswaldo Cruz, Av. Brasil, 4365 - Manguinhos, Rio de Janeiro - RJ, 21040-900, Brazil; Laboratório de Micobactérias, Instituto de Microbiologia Paulo de Góes, Universidade Federal do Rio de Janeiro, Av. Carlos Chagas Filho, 373 - bloco I - Cidade Universitária, Rio de Janeiro - RJ, 21941-970, Brazil; Laboratório de Informação em Saúde, Instituto de Comunicação e Informação Científica e Tecnológica em Saúde, Fundação Oswaldo Cruz, Av. Brasil, 4365 – Manguinhos, Rio de Janeiro – RJ, 21040-900, Brazil

**Keywords:** COVID-19, Tuberculosis, SARS-CoV-2, Coinfection

## Abstract

Due to tuberculosis (TB) patients’ pulmonary damages, some authors believe that a SARS-CoV-2 coinfection may result in unfavorable outcomes. A cross-sectional anti-SARS-CoV-2 antibodies seroprevalence study was conducted at a TB treatment tertiary referral unit in Rio de Janeiro, Brazil, to estimate the proportion (in %) of TB patients exposed to the new coronavirus and their main outcomes. Of 83 patients undergoing TB treatment, 26.5% have already been infected by the new coronavirus. Most patients were asymptomatic (69%) or had mild COVID-19 cases (31%). Only one patient required hospitalization. Among the symptoms and signs presented, the most frequently reported were: fever, headache, and myalgia. People with less education and less purchasing power seemed to had been more exposed to SARS-CoV-2.

## 1. Introduction

COVID-19 (Coronavirus Disease 2019), caused by the new Betacoronavirus SARS-CoV-2, was first described in Wuhan, China, in December 2019, as responsible for severe pneumonia [1], [2], among several other systemic and organ specific manifestations [3]. Despite the measures taken to curb the epidemic, SARS-CoV-2 has spread throughout the world and has caused thousands of deaths. Until July 30, 2021, 196.794.025 cases and 4.202.385 deaths by COVID-19 had been confirmed worldwide. As of the same date, Brazil had 19.839.369 confirmed cases and 554.497 deaths [4]. In contrast to the newest infectious disease to be defined as a pandemic, tuberculosis (TB) is an ancient disease caused by *Mycobacterium tuberculosis* [5]. TB mainly affects the lungs and, until the COVID-19 emergence, it was the leading death caused by a single infectious agent in the world, with estimates, in 2018, of approximately 10 million cases and 1.5 million deaths [6].

Due to TB patients’ pulmonary functional impairment and unfavorable outcomes in consequence of putative associations with fungal, viral and bacterial pulmonary infections [7]–[11], it has been suggested that TB and COVID-19 coinfection could result in more severe conditions and higher mortality rates [12]. Despite many published studies on COVID-19 in recent months, few have analyzed how this new disease may affect TB patients.

Seroprevalence studies are essential because, in addition to estimating the proportion of the population previously infected, helping to assess the time required to reach herd immunity, it may also help to better understand disease transmission patterns [13]. Therefore, to plan an adequate public health response to tuberculosis patients in pandemic times, ideally discerning and anticipating the present and future disease dynamic, the anti-SARS-CoV-2 antibodies level may be crucial. Although some antibody seroprevalence studies for SARS-CoV-2 have been published [13]–[18], none have focused on TB patients (to the best of our knowledge). Thus, we aimed to estimate the seroprevalence of SARS-CoV-2 antibodies in patients undergoing TB treatment and investigate the demographic and clinical characteristics of this population exposed to the new coronavirus.

## 2. Methods

### 2.2 Study design

A cross-sectional anti-SARS-CoV-2 antibodies seroprevalence study was carried out among a convenience sample of TB and drug-resistant TB (DR-TB) patients assisted at a tertiary referral unit in Rio de Janeiro, Brazil. Participants were screened between September 01, 2020 up to February 28, 2021.

### 2.2 Study Subjects

The *Centro de Referência Professor Hélio Fraga* (CRPHF) is a tertiary reference health unit for TB located in Rio de Janeiro city, Rio de Janeiro, Brazil. Rio de Janeiro is the capital of the homonymous Brazilian state, which has reported the highest new case numbers of TB in 2019 (93.7 cases per 100,000 inhabitants) [6]. Rio de Janeiro was also one of the Brazilian cities with the highest COVID-19 number in 2020, couting 215,016 confirmed [19].

The CRPHF outpatient clinic treats patients with DR-TB, including multidrug-resistant TB (MDR-TB), extensively drug-resistant TB (XDR-TB), and TB special cases (patients with susceptible-TB who need different treatments due to basic regimen adverse effects). The present study included patients under medical care at the CRPHF undergoing treatment for TB or TB-DR, aged 18 years old or more.

### 2.3 Data collection

Participants answered short standard questionnaires, including socio-demographic information, symptoms related to COVID-19, and access to COVID-19 diagnostic tests. Medical records were accessed to investigate clinical and therapeutic TB information.

### 2.4 SARS-CoV-2 serological test

Antibodies prevalence was assessed by the rapid test TR DPP^®^ COVID-19 IgM/IgG (Bio-Manguinhos/Fiocruz, Rio de Janeiro, Brazil), using one drop of blood from a finger prick sample. It is an immunochromatographic test on a differentiated double-path platform for the qualitative detection of IgM and IgG antibodies anti-SARS-CoV-2 virus. An electronic device (Micro Reader DPP®) was used to assist in the interpretation of results, reactive or non-reactive.

The TR DPP^®^ COVID-19 IgM/IgG (Bio-Manguinhos) was licensed by the Brazilian Health Surveillance Agency (ANVISA) in April 2020 [20]. According to the manufacture specifications its sensitivity was estimated as 79% (95%CI:70.9-86.8%) for IgM and 95% (95%CI:88.8-97.9%) for IgG. The specificity for IgM was estimated as 98.0% (95%CI:95.8-99%) and 97% (95%CI:94-98%) for IgG.

### 2.5 SARS-CoV-2 Real time RT-PCR (RT-qPCR)

The SARS-CoV-2 RT-qPCR was performed for all patients. Naso and oropharyngeal secretions were collected and subjected to total RNA extraction using QIAmp Viral RNA Mini kit (Qiagen, Hilden, Germany). SARS-CoV-2 detection was carried out by SARS-CoV-2 RT-qPCR using the SARS-CoV-2 Molecular E/RP kit (Biomanguinhos, Rio de Janeiro, Brazil) based on the protocol previously designed by Corman et al. [21]. The analyzes were performed at the Respiratory and Measles Viruses Laboratory of the Oswaldo Cruz Institute (IOC/Fiocruz - SARS-CoV-2 National Reference Laboratory for the Brazilian Ministry of Health).

### 2.6 Data analyses

The enrolled patients were characterized using descriptive statistics, stratified by anti-SARS-CoV-2 antibody results. Groups were compared using Fisher’s exact test for categorical variables. Anti-SARS-CoV-2 antibodies prevalence and 95% CIs were calculated as the number of positive test results divided by the total number of participants.

Data were collected in electronic forms organized on *FormSus* version 3.0 (DATASUS), and all analyses were performed using R version 4.1.1 and R Studio version 1.2.1335 software (R Foundation for Statistical Computing).

### 2.7 Ethical considerations

This study was reviewed and approved by the research ethics committee of the Sergio Arouca National School of Public Health / Oswaldo Cruz Foundation (CAAE: 32219620.9.0000.5240). Written informed consent was obtained from all study participants.

## 3. Results

A total of 83 participants were interviewed and tested for SARS-CoV-2. Overall study participants were male (60%), 30-59 age group (65%), brown (46%) and 72% with eight or more years of schooling. 39% of participants lived in impoverished communities and 58% declared have a monthly income between one and three minimum wages (Brazilian minimum wages - R$ 1,100, about $219 [at the time of their assessment]). Only 14% of participants reported having been previously tested by RT-qPCR for COVID-19 diagnosis, while 25% reported having undergone rapid serological testing. Furthermore, 4.8% stated having had COVID-19 previously. About TB, most participants had pulmonary TB (88%), 70% were new cases and 73% were DR-TB, of which 33% were MDR-TB. The baseline population study characteristics are depicted in **Table 1**.

**Table 1.**
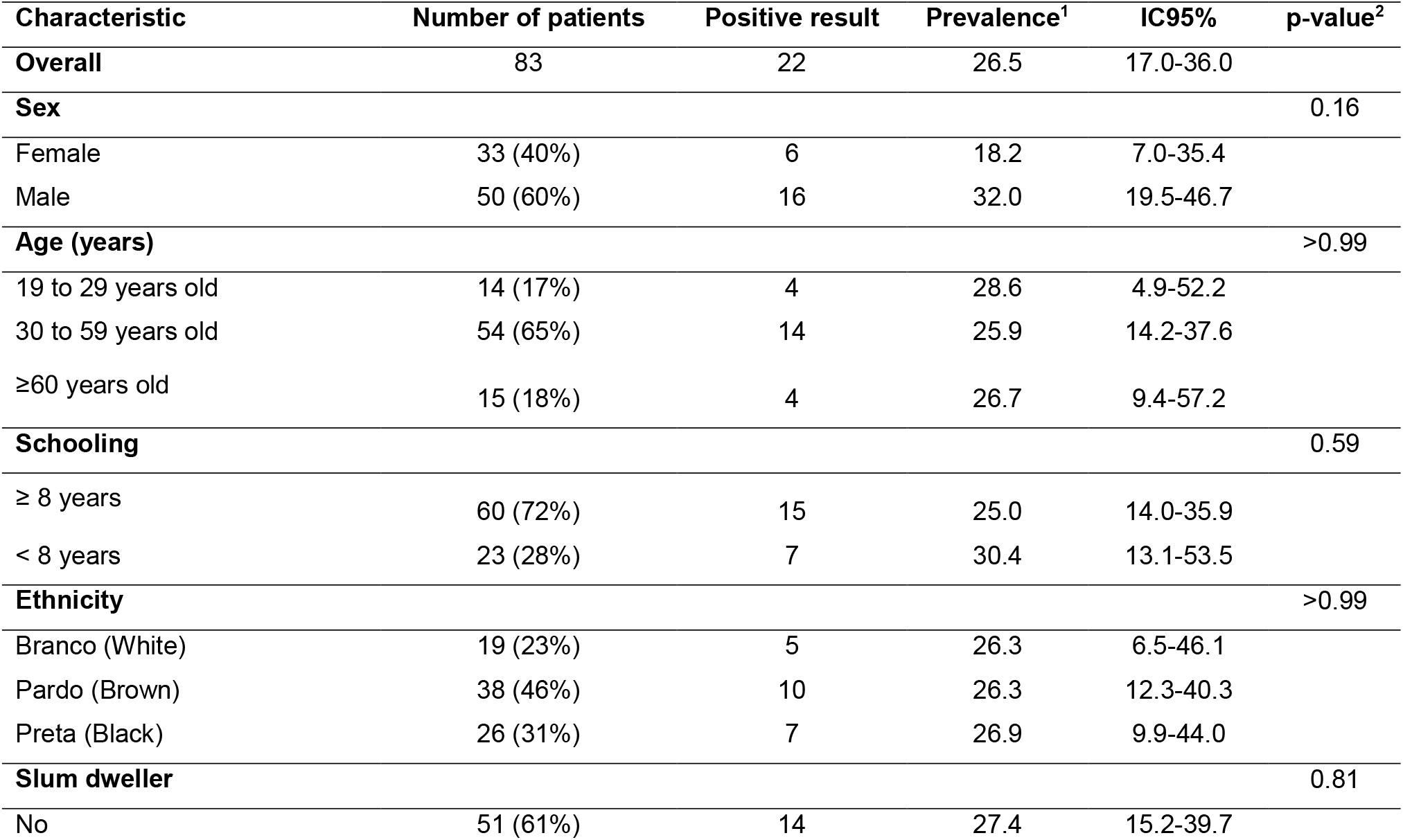
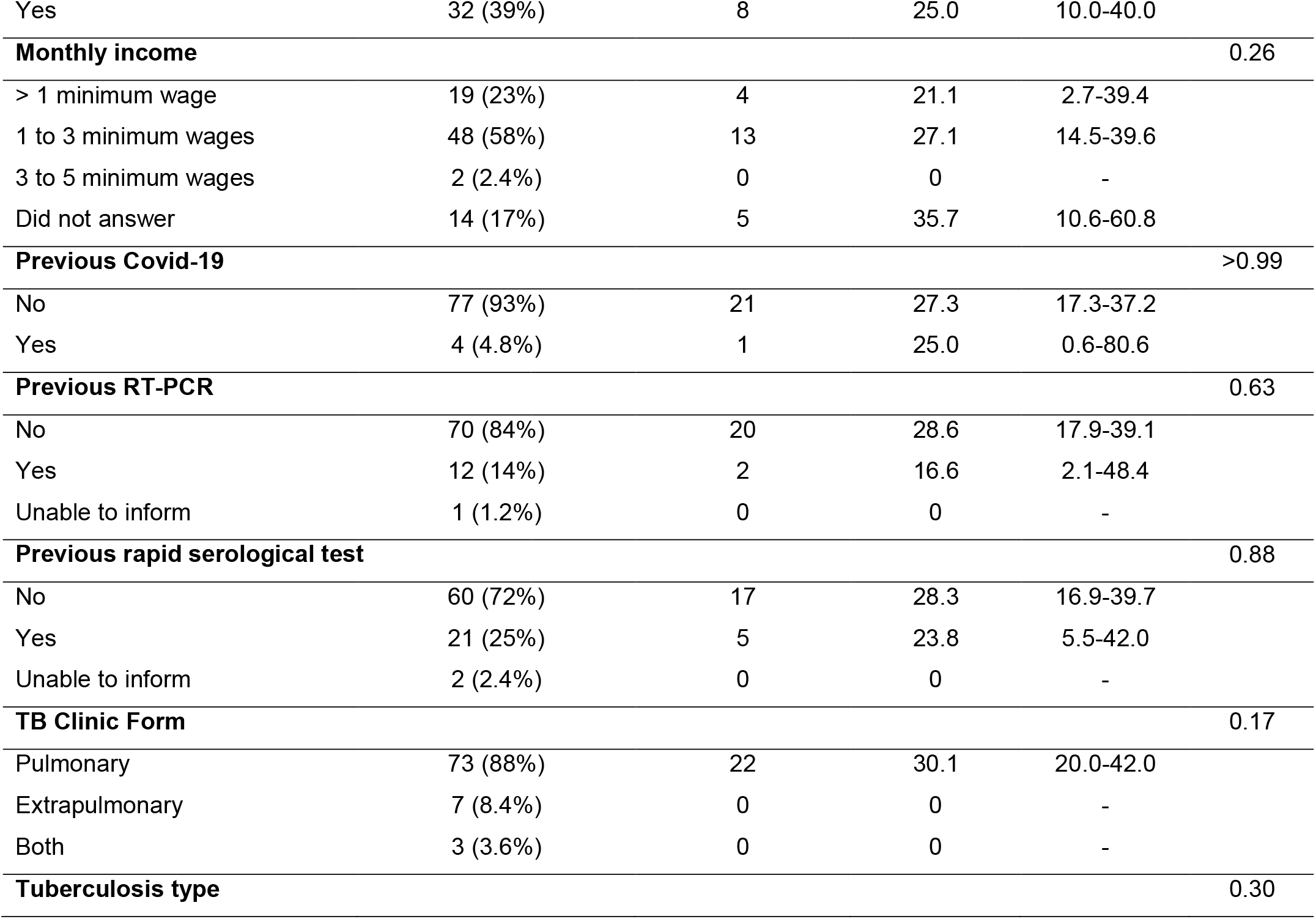
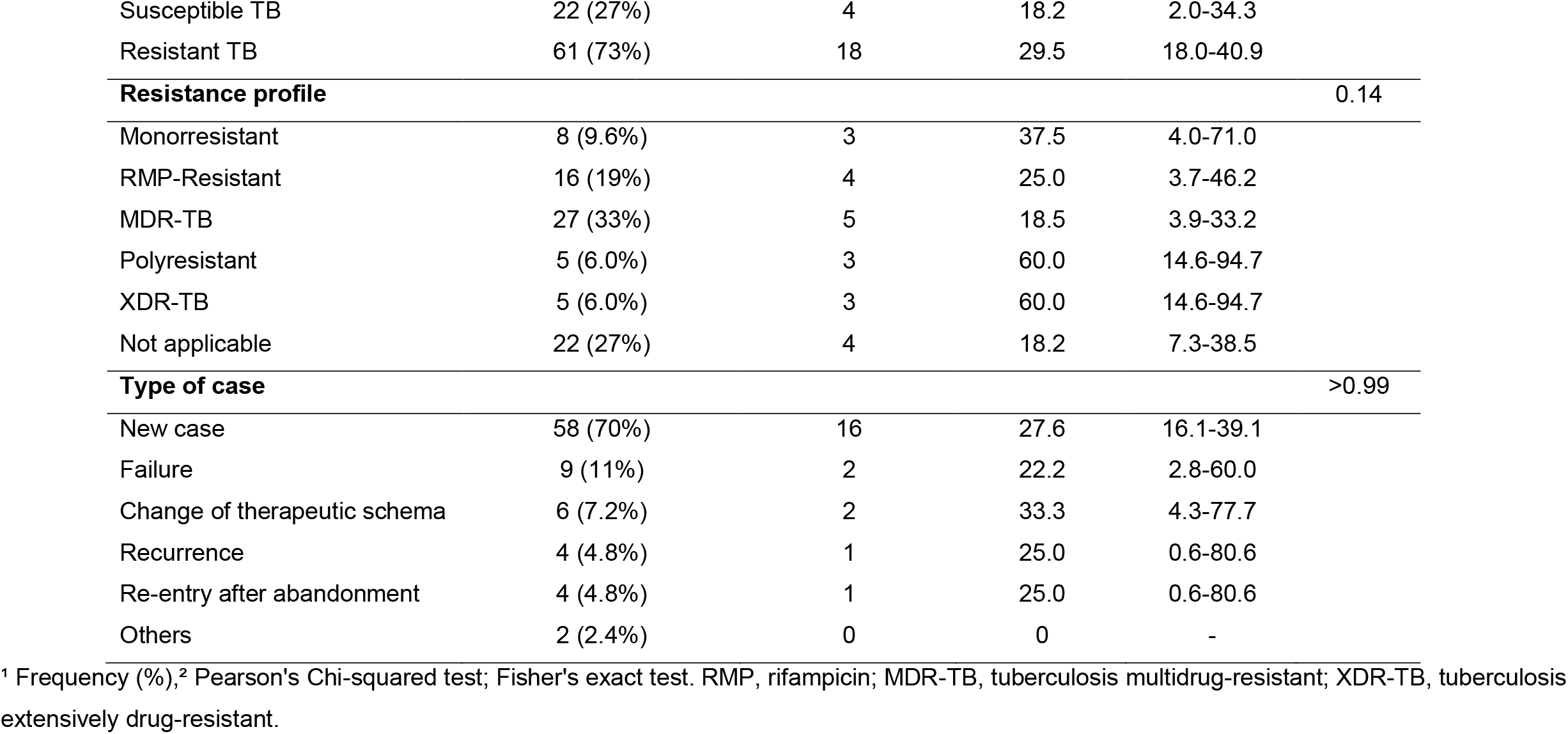
SARS-CoV-2 seroprevalence, sociodemographic indicators, previous tests and tuberculosis profile of patients under tuberculosis treatment assisted by a tertiary referral unit in Rio de Janeiro, Brazil.

A total of 22 (26.5%; 95%CI:17.0–36.0%) individuals were seropositive for anti-SARS-CoV-2 antibodies. The prevalence by demographics and clinical characteristics is shown in **Table 1**, highlighting: 28.6% (CI95%:4.9-52.2) for people between 19 to 29 years old, 30.4% (CI95%:13.1-53.5) for people with less than eight years of schooling, 25.0% (CI95%:10.0-40.0) people living in slums, and 27.1% (CI95%:14.5-39.6) patients that receive monthly income between one and three salaries. SARS-CoV-2 antibodies seropositive results were only observed in patients with pulmonary TB (seroprevalence 30.1%; CI95%:20.0-42.0). For individuals with DR-TB, the point seroprevalence was 29.5% (95%CI:18.0-40.9) whereas for susceptible TB was 18.2% (CI95%:2.0-34.3).

By RT-PCR, SARS-CoV-2 was detected in three patients (**Table 2**: patients 2, 7 and 18). Patients 2 and 18 non-reagents in the serological test, while patient 7 seropositive. Four patients reported having been previously COVID-19 diagnosed (**Table 2**: patients 2, 5, 22 and 23). Patients 2 and 5, the diagnostic test has been performed approximately 30 days before the interview. Patient 2 presented positive RT-PCR and a negative serological test. Patient 5 presented only a positive serological test. Patients 22 and 23, the diagnostic test has been performed nine months before the enrollment in the study and both presented negative results of either RT-PCR and serological test.

**Table 2.** Tuberculosis profile and COVID-19 test results of diagnosed patients and/or patients with SARS-CoV-2 positive serology.

|  | **Clinical Form** | **TB Resistance Profile** | **Prior TB treatment** | **COVID-19 Previous Diagnosis** | **Serological Test Result** | **RT-qPCR Result (Ct)\*** |
| --- | --- | --- | --- | --- | --- | --- |
| 1 | Pulmonary | Monoresistant | 1 | No | IgM and IgG | Non-detected |
| 2 | Pulmonary | RMP Resistant | No | 1 month before the interview | Negative | Detected (20,7) |
| 3 | Pulmonary | MDR-TB | 1 | No | IgG | Non-detected |
| 4 | Pulmonary | Polyresistant | 1 | No | IgM and IgG | Non-detected |
| 5 | Pulmonary | Monoresistant | No | 1 month before the interview | IgM and IgG | Non-detected |
| 6 | Pulmonary | Polyresistant | 2 | No | IgG | Non-detected |
| 7 | Pulmonary | MDR-TB | 1 | No | IgM and IgG | Detected (34,9) |
| 8 | Pulmonary | RMP Resistant | 1 | No | IgM and IgG | Non-detected |
| 9 | Pulmonary | Monoresistant | 2 | No | IgM | Non-detected |
| 10 | Pulmonary | MDR-TB | 1 | No | IgG | Non-detected |
| 11 | Pulmonary | RMP Resistant | No | No | IgG | Non-detected |
| 12 | Pulmonary | MDR-TB | 1 | No | IgG | Non-detected |
| 13 | Pulmonary | Susceptible | 1 | No | IgG | Non-detected |
| 14 | Pulmonary | Susceptible | No | No | IgG | Non-detected |
| 15 | Pulmonary | XDR-TB | No | No | IgG | Non-detected |
| 16 | Pulmonary | Polyresistant | 1 | No | IgM | Non-detected |
| 17 | Pulmonary | RMP Resistant | No | No | IgM | Non-detected |
| 18 | Pulmonary | Susceptible | No | No | Negative | Detected (24,0) |
| 19 | Pulmonary | Susceptible | No | No | IgG | Non-detected |
| 20 | Pulmonary | RMP Resistant | 1 | No | IgG | Non-detected |
| 21 | Pulmonary | XDR-TB | 10 | No | IgG | Non-detected |
| 22** | Extrapulmonary | Susceptible | No | 9 months before the interview | Negative | Non-detected |
| 23 | Pulmonary | Susceptible | 1 | 9 months before the interview | Negative | Non-detected |
| 24 | Pulmonary | Susceptible | No | No | IgG | Non-detected |
| 25 | Pulmonary | XDR-TB | 2 | No | IgM and IgG | Non-detected |
| 26 | Pulmonary | MDR-TB | No | No | IgG | Non-detected |

\* Ct values for SARS-CoV-2 target “E”; \*\*Patient who required hospitalization. RMP, rifampicin; MDR-TB, tuberculosis multidrug-resistant; XDR-TB, tuberculosis extensively drug-resistant.

Adding to patients with a positive serology for SARS-CoV-2, those who detection by RT-PCR and had been previously diagnosed with COVID-19, there were 26 positive patients, 69% (18/26) of them asymptomatic. Among the 31% (8/26) who had had symptoms, 50% reported having up to 4 signs and the other half declared to have had from five to ten. The most common symptoms reported were fever, headache, and myalgias, each one with 75% of reports (multiple answers allowed) (**Table 3**).

**Table 3.**
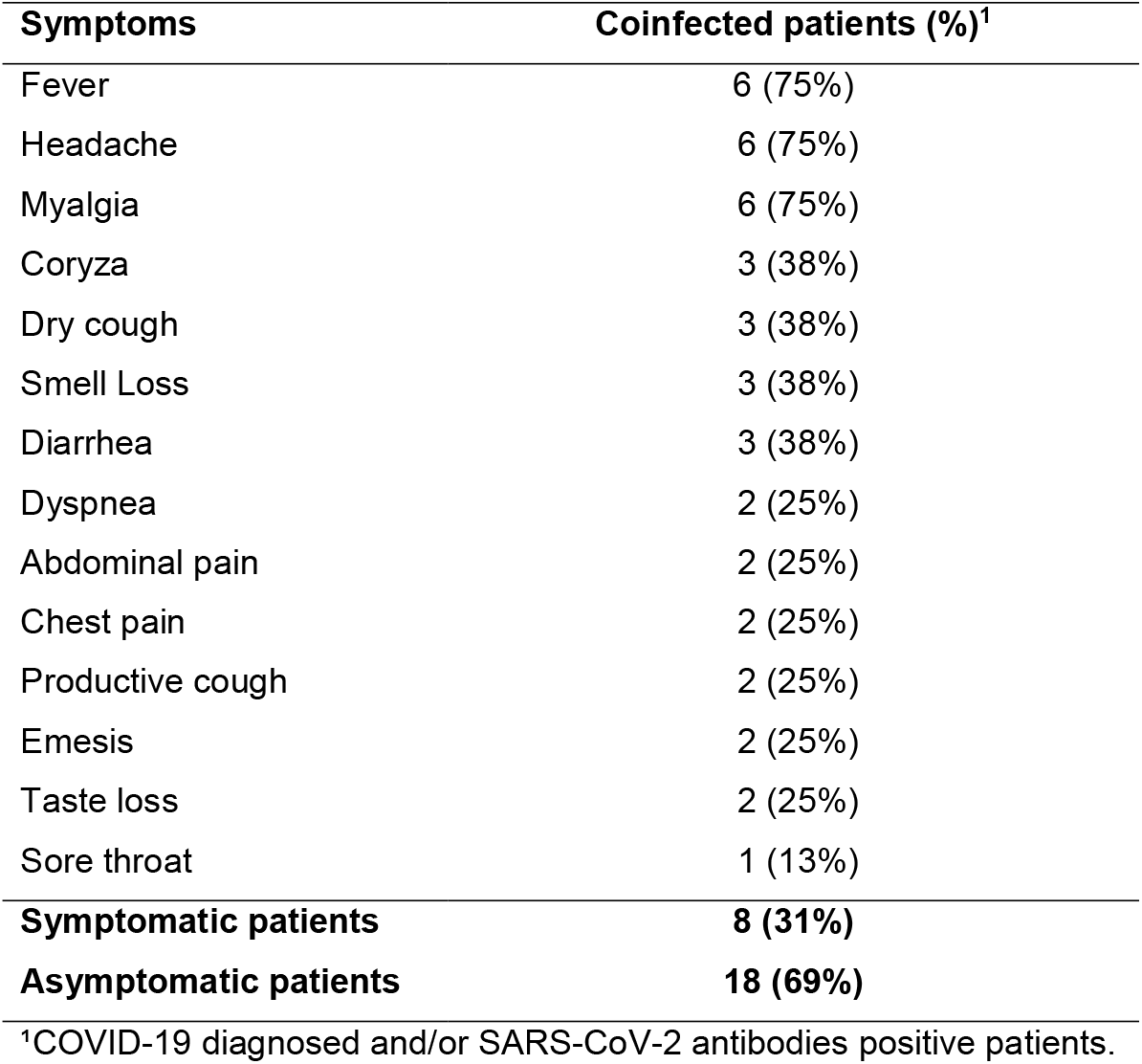
Symptoms reported by patients coinfected with TB and SARS-CoV-2 assisted by a tertiary referral unit in Rio de Janeiro, Brazil.

| Symptoms | Coinfected patients (%)<sup>1</sup> |
| --- | --- |
| Fever | 6 (75%) |
| Headache | 6 (75%) |
| Myalgia | 6 (75%) |
| Coryza | 3 (38%) |
| Dry cough | 3 (38%) |
| Smell Loss | 3 (38%) |
| Diarrhea | 3 (38%) |
| Dyspnea | 2 (25%) |
| Abdominal pain | 2 (25%) |
| Chest pain | 2 (25%) |
| Productive cough | 2 (25%) |
| Emesis | 2 (25%) |
| Taste loss | 2 (25%) |
| Sore throat | 1 (13%) |
| **Symptomatic patients** | **8 (31%)** |
| **Asymptomatic patients** | **18 (69%)** |

<sup>1</sup>COVID-19 diagnosed and/or SARS-CoV-2 antibodies positive patients.

Among all patients included, 53% (44/83) reported belonging to some priori defined risk group for COVID-19, the most frequently reported were: to be aged over 60 years old (32%) and diabetes (27%). Among patients have been infected by SARS-CoV-2, 42% (11/26) presented characteristics associated with a poor prognosis, and the most frequently reported were to be aged over 60 years (15%) and diabetes mellitus (12%) (**Table 4**).

**Table 4.** Risk factors for COVID-19 reported by patients under TB treatment assisted by a tertiary referral unit in Rio de Janeiro, Brazil.

| **COVID-19 Risk factors** | **Total** | **Coinfected patients<sup>1</sup>** |
| --- | --- | --- |
| Over 60 years old | 14 (32%) | 4 (15%) |
| Pregnant | 1 (2.3%) | 0 (0%) |
| Heart disease | 1 (2.3%) | 0 (0%) |
| Hypertension | 3 (6.8%) | 1 (4%) |
| Diabetes | 12 (27%) | 3 (12%) |
| Kidney disease | 3 (6.8%) | 1 (4%) |
| Neurological disease | 2 (4.5%) | 1 (4%) |
| Cancer | 1 (2.3%) | 0 (0%) |
| Smoking | 6 (14%) | 1 (4%) |
| HIV | 6 (14%) | 2 (8%) |
| **Total patients with risk factors** | **44 (53%)** | **11 (42%)** |
| **Total patients without risk factors** | **35 (42%)** | **12 (46%)** |
| **Did not know or want to inform** | **4 (5%)** | **3 (12%)** |

<sup>1</sup>COVID-19 diagnosed and/or SARS-CoV-2 antibodies positive patients.

Only one patient needed hospitalization because of COVID-19. This patient was previously COVID-19 diagnosed, and in the present study presented negative results for both RT-PCR and serology tests. This patient was diagnosed with TB after has presented COVID-19 symptoms (**Table 2**). This case was the most severe reported in the context of the present study.

## 4. Discussion

This is the first Brazilian anti-SARS-CoV-2 antibodies seroprevalence study in TB patients. The comprehensive assessment of seroprevalence is key because it provides information about the extent of transmission in the past and may help to predict the future course of the pandemic [13], [14]. When this kind of study targets a specific segment of the population, we may infer how these people have been affected. In the present case, the research focus was patients under TB and DR-TB treatment. In addition to COVID-19 tests, a questionnaire was also answered by the patients to assess putative associations and to better understand how these patients have been affected by the COVID-19 pandemic.

Our results showed that 26.5% of patients undergoing TB treatment had been infected with the new coronavirus. However, the seroprevalence of anti-SARS-CoV-2 antibodies in the present study was higher than previously reported for the city of Rio de Janeiro until now, 7.5% (95%CI:4.5–11.7). This result was reported in a study carried out between April and June 2020 and refers to the general population of Rio de Janeiro [15]. The authors also observed an increase of more than three times between surveys performed 2 to 3 weeks apart (2.4% vs 7.5%). Between June 2020 (last month evaluated by Hallal et al. [15]) and February 2021 (last month when patients were enrolled in the present study), the COVID-19 confirmed cases in the city increased from 56,936 to 188,833 cases [22]. Therefore, the seroprevalence differential proportion observed when this study is cross-compared with previous papers may be reflecting the increment in the number of cases that occurred during this period. Furthermore, the fact these different studies have been nested in totally different sampling strategies may help to better understand such marked discrepancies [23]. Several other factors may explain the observed differences, such as different timing, the different tests used in the studies, as well as intrinsic differences between patients recruited in communities compared to those attending health services.

Only one former anti-SARS-CoV-2 seroprevalence study assessed TB cases. This research was carried out in Cape Town, South Africa, from August to September 2020. They showed that 26.7% of the subjects who reported previous TB had positive serology for SARS-CoV-2, while for the general population it was 23.7% [18]. Gao et al. [24], in a meta-analysis of six studies conducted in China, identified a prevalence of TB among patients with COVID-19 ranging from 0.47 to 4.47%. These authors found that a higher TB prevalence was identified among patients with severe COVID-19 than among non-severe cases (1.47%, 10/680 vs 0.59%, 10/1703; OR: 2.1; p-value=0.24). We must keep in mind that this meta-analysis refers to a set of “reverse” studies, i.e. studies assessing TB among patients with COVID-19, instead of assessing SARS-CoV-2 among patients under management and care for TB.

In addition to the small number of publications focused on TB and SARS-CoV-2 coinfection, published studies have presented controversial data. While some point to more severe outcomes and even a higher mortality rate among TB patients infected with the new coronavirus [24]–[26], others found a weak or no association between disease severity and/or death in TB patients diagnosed with COVID-19 [27]–[29].

In the present study, most patients had mild COVID-19 cases. Among patients with positive serology or a COVID-19 confirmed diagnosis, 69% were asymptomatic, a percentage higher than the ∼30% estimated by previous studies [30]–[34]. Among symptomatic patients, fever, headache and myalgia were the most frequently signs reported, differing somewhat from the most common symptoms reported in other studies (fever, dry cough, and dyspnea) [25], [29], [35]. This difference may be related to the fact our patients presented milder COVID-19 cases vis-à-vis those from the abovementioned studies.

Although most people develop specific antibodies against SARS-CoV-2 after infection, patients with milder COVID-19 symptoms seem to produce smaller amount of antibodies [36]. In addition, some works have showed a decline of anti-SARS-CoV-2 antibody titers over time [36]–[38]. This information may help to explain the seronegative results of two patients who have been COVID-19 diagnosed nine months before the interview. Furthermore, the detection limit of the test and the time required to seroconversion (that may varies among patients [38]), may also interfere with anti-SARS-CoV-2 antibodies detection. These are possible reasons for what might have happened to the patient who had been COVID-19 diagnosed one month before the interview and had no detected antibodies. Due to all abovementioned reasons, we cannot exclude a previous infection in patients with negative serology for SARS-CoV-2.

It was impossible to infer any important correlation between the Ct value and serologic response among the patients tested, since only three subjects presented RT-PCR positive results. However, it is worthy to note that the patient which presented antibodies against the virus was the one with the highest Ct value (Ct= 34,9). Such result reinforces the well-known disconnection between virus load and humoral immune response of the patient during respiratory viral infections as observed in other diseases like influenza and syncytial respiratory virus infections.

In our study, only one patient required hospitalization, a patient diagnosed with extrapulmonary TB 8 months after the COVID-19 symptoms. TB diagnosis after SARS-CoV-2 infection has been reported by Tadolini et al. [35]. More extensive studies are needed to understand any role of SARS-CoV-2 in the progression of TB infection to disease.

Although our small sample size did not allow us to observe significant differences among the socio-demographic categories (putative beta error), some findings should be highlighted. While 72% of respondents reported having more than 8 years of schooling, people with less education seem to be more exposed to SARS CoV-2, as well as those with less purchasing power. This data agrees with results by Horta et al. [16], who analyzed socioeconomic and ethnic status in a seroprevalence study that gathered samples from different Brazilian states.

The high DR-TB rate found in our study seems to be associated to the fact that the approach took place in a tertiary health unit, where patients with DR-TB have been mainly referred to management and care. Some authors have questioned whether DR-TB cases would be an additional risk factor for COVID-19 [28]. In the present study, most 73% (61/83) coinfected patients had TB with some resistance, and, in addition, more than half of coinfected patients have undergone at least one previous TB treatment. Even with these potential aggravating factors, they were asymptomatic or had mild cases of COVID-19.

An important measure to curb the COVID-19 spread is mass testing. Unfortunately, this vital strategy to control the pandemic seems has not been carried out in Rio de Janeiro. In our study, 84% of respondents did not have access to RT-PCR tests and 72% to serological tests.

This study has some limitations. First, the possible selection bias, since the sample in this study included those who naturally tend to be more mobile and less likely to keep social distancing, first of all to attend their appointments at health facilities, but also to address their bare necessities since most of them are poor and live in impoverished and underserved communities. Second, the small sample size, notwithstanding corresponding to the caseload of a tertiary, referral, health service, are likely affected by type II errors. Furthermore, questionnaires, especially when applied to studies dealing with long-term patients, living with severe diseases, tend to be affected by both recall [39] and social desirability bias [40].

## 5. Conclusions

The present study sought to estimate the prevalence of COVID-19 in patients with TB treatment in Rio de Janeiro, Brazil, considering the fact that this vulnerable population is likely to be severely affected by the epidemic. The 26.5% rate of anti-SARS-CoV-2 antibodies in a population with severe lung problems is of concern, although the risk of serious complications and death remains controversial.

Knowing the anti-SARS-CoV-2 antibodies prevalence among patients is essential to better understand COVID-19 spread and help policymakers to implement evidence-based policies to curb the epidemic and to provide coinfected patients proper management and care. Our data suggest that the high seroprevalence observed among TB patients may be closely associated to the low socioeconomic level of these patients, besides their pressing need to attend successive appointments to manage and care their underlying and putative new medical conditions. To fully adopt social distancing measures in this context, despite being essential, remains a major challenge. In addition to the systematic use of personal protective equipment, these patients should be prioritized by vaccination efforts.

## Data Availability

All data produced in the present study are available upon reasonable request to the authors

## Acknowledgements

To all team members at the Germano Gerhardt Research Outpatient Clinic of the Centro de Referência Professor Hélio Fraga for their support during the development of this project. Bio-Manguinhos (Fiocruz) for the rapid test kits TR DPP® COVID-19 IgM/IgG donation. To the Respiratory Virus and Measles Laboratory (IOC/Fiocruz) for carrying out the SARS-CoV-2 qRT-PCR tests.

## Funding

This research did not receive any specific grant from funding agencies in the public, commercial, or not-for-profit sectors.

## CRediT author statement

**Karen Machado Gomes:** Conceptualization, Methodology, Investigation, Data Curation, Writing -Original Draft, Review & Editing, Visualization, Supervision. **Suzanne Pereira Leite:** Methodology, Investigation; Resources; Supervision. **Maria Helena Vieira Moutinho:** Investigation. **Thatiana Alfena de Souza:** Investigation. **Rita de Cássia Batista:** Investigation. **Luiz Claudio Motta de Oliveira:** Investigation. **Paulo Redner:** Investigation. Writing - Review. **Jesus Pais Ramos:** Writing - Review. **Fatima Maria Gomes da Rocha:** Resources. **Gisele Pinto de Oliveira:** Resources. **Antônio Teva:** Resources, Writing - Review. **Fernando do Couto Motta:** Investigation. **Marilda Agudo Mendonça Teixeira de Siqueira:** Resources. **Rafael Silva Duarte:** Writing - Review. **Francisco Inácio Pinkusfeld Monteiro Bastos**: Writing – Review. **Paulo Victor de Sousa Viana:** Methodology, Formal analysis, Investigation, Resources, Data Curation, Writing Original Draft, Review & Editing.

## Declaration of interest

None

## Notes

### Competing Interest Statement

The authors have declared no competing interest.

### Funding Statement

This study did not receive any funding.

